# Interplay of antibody and cytokine production reveals CXCL-13 as a potential novel biomarker of lethal SARS-CoV-2 infection

**DOI:** 10.1101/2020.08.24.20180877

**Authors:** Alexander M. Horspool, Theodore Kieffer, Brynnan P. Russ, Megan A. DeJong, M. Allison Wolf, Jacqueline M. Karakiozis, Brice J. Hickey, Paolo Fagone, Danyel H. Tacker, Justin R. Bevere, Ivan Martinez, Mariette Barbier, Peter L. Perrotta, F. Heath Damron

## Abstract

The SARS-CoV-2 pandemic is continuing to impact the global population. This study was designed to assess the interplay of antibodies with the systemic cytokine response in SARS-CoV-2 patients. We demonstrate that significant anti-SARS-CoV-2 antibody production to Receptor Binding Domain (RBD), Nucleocapsid (N), and Spike S1 subunit (S1) of SARS-CoV-2 develops over the first 10 to 20 days of infection. The majority of patients produced antibodies against all three antigens (219/255 SARS-CoV-2 positive patient specimens, 86%) suggesting a broad response to viral proteins. Patient mortality, sex, blood type, and age were all associated with differences in antibody production to SARS-CoV-2 antigens which may help explain variation in immunity between these populations. To better understand the systemic immune response, we analyzed the production of 20 cytokines by SARS-CoV-2 patients over the course of infection. Cytokine analysis of SARS-CoV-2 positive patients exhibited increases in proinflammatory markers (IL-6, IL-8, IL-18) and chemotactic markers (IP-10, SDF-1, MIP-1*β*, MCP-1, and eotaxin) relative to healthy individuals. Patients who succumbed to infection produced decreased IL-2, IL-4, IL-12, IL-13, RANTES, TNF-α, GRO-α, and MIP-1α relative to patients who survived infection. We also observed that the chemokine CXCL13 was particularly elevated in patients that succumbed to infection. CXCL13 is involved in B cell activation, germinal center development, and antibody maturation, and we observed that CXCL13 levels in blood trended with anti-SARS-CoV-2 antibody production. Furthermore, patients that succumbed to infection produced high CXCL13 and also tended to have high ratio of nucleocapsid to RBD antibodies. This study provides insights into SARS-CoV-2 immunity implicating the magnitude and specificity of response in relation to patient outcomes.

## Introduction

The SARS-CoV-2 pandemic has drastically affected life in the United States and across the globe. As of August 2020, more than 5.6 million people in the United States have been infected and over 175,000 patients have died^1^. SARS-CoV-2 rapidly infected urban centers in California, New York, and other major cities across the United States that until recently were the main source of United States SARS-CoV-2 infections. Studies of anti-SARS-CoV-2 published in the first months of the pandemic were highly focused, with little exploration of the broader immune response in COVID-19 patients. Since then, several factors including elevated pro-inflammatory cytokines and others have been identified in SARS-CoV-2 pathology^2–7^ but little is known about the interplay between cytokine production and the antibody response during SARS-CoV-2 infection. CXCL13 is a cytokine integral to germinal center formation^8-10^ and has been used as a biomarker of an anti-infective antibody response^8^. B cells are attracted to the germinal center via production of CXCL13^11,12^ by follicular dendritic cells and T follicular helper cells^13,14^. Production of CXCL13 and B cell germinal center formation promotes somatic hypermutation and affinity maturation of antibodies with virus-neutralizing function^8^. CXCL13 production is quantifiable in human serum^15^, and has not been characterized in the context of coronavirus infection in humans. In this respect, we sought to characterize the interplay of the antibody-mediated immunological response and the cytokine-mediated response to SARS-CoV-2 infection with a focus on CXCL13. We studied well-known markers of anti-viral immunity including: antibody production to the SARS-CoV-2 receptor-binding domain (RBD), nucleocapsid (N), and spike s1 (S1) protein domains, Th1 and Th2 associated cytokine production, and CXCL13 production to characterize the immune profile of COVID-19 patients. Our study provides a broad view of the anti-SARS-CoV-2 immune response and reveals that CXCL13 may serve as a novel predictor of lethal infection in COVID-19 patients.

## Methods

### Patient sampling and analysis

Clinical results of molecular FDA emergency use authorization (EUA) approved assays (Abbott M2000, BD Max, Cepheid GeneXpert) were reviewed by querying an electronic health record (HER, Epic, Vernona, WI) at least 3 times a week, for admitted in-patients with positive or negative COVID-19/SARS-CoV-2 test results at West Virginia University Hospital (WVUH) (see all patient information in Supplementary Data File). All available residual serum and plasma clinical specimens (492 from 82 inpatients, collected from day 0 to day 55 post symptom onset) were then retrieved, de-identified, and stored at −80°C. Electronic medical records were reviewed (IRB #2004976401) for symptoms, date of symptom onset, and patient demographic information (age, sex, mortality). If no symptoms were recorded, the date of admittance was documented as date of symptom onset. Serological results from EUA approved antibody testing (Abbott Architect) performed in the WVUH clinical laboratory and ABO blood type were documented when available. Patient specimens were de-identified by appropriately CITI trained clinical research staff before transfer to the WVU Vaccine Development center research laboratory for testing and aliquoting. Once specimens were thawed for testing, residual specimens were aliquoted into 1 mL cryovials in 0.5 mL increments and frozen at −80°C. No specimens underwent >3 free thaw cycles prior to testing to prevent degradation.

### Production of SARS-CoV-2 RBD protein

Production of the SARS-CoV-2 RBD protein was done by transient transfection of the HEK293T cells with a pCAGGS mammalian expression vector containing an RBD construct with a C-terminal hexa-histidine tag and codon optimized for mammalian expression (pCAGGS vector catalog number NR-52309, BEI Resources, Manassas, VA, USA). SARS-CoV-2 RBD protein was produced by transient transfection of HEK293T cells cultured in 300 cm^2^ flasks. Each flask with 60-80% confluent cells was transfected with 60 μg of plasmid DNA complexed with 120 μg of 25 kDa linear polyethylenimine (PEI)(Polysciences Inc., Warrington, PA, USA). The DNA/PEI complex was prepared by slowly adding the PEI solution (0.08 mg/mL in PBS) to the DNA solution (0.04 mg/mL in PBS) with continuous mixing followed by 10 minutes incubation at room temperature; the DNA/PEI complex was then diluted with 45 mL of serum-free DMEM medium and used to replace the FBS supplemented DMEM medium in the flask. For the production of the SARS-CoV-2 N protein, HEK293T cells were harvested 72 hours post transfection and kept at −8°C until processing. For the production of the SARS-CoV-2 RBD protein, which is secreted into the culture medium, the cell culture medium was collected after 48 hours, stored at 4°C, and replaced with fresh serum-free DMEM medium; after an additional 48 hour, corresponding to 96 hours post transfection, the medium was collected, pooled with the 48 hour post transfection medium and stored at 4°C until processing. Five hundred mL of medium were supplemented with 500 U of Pierce Universal Nuclease (Thermo Fisher Scientific), centrifuged at 4000 × *g* for 20 minutes and the supernatant filtered through a 0.45 μm PES membrane. The filtered medium was applied onto 5 mL HisTrap FF cartridges (GE Healthcare Bio-Sciences) installed on an AKTA Purifier run with buffer A (20 mM NaH_2_PO_4_, 0.5 M NaCl, 10 mM Imidazole, pH 7.4) and buffer B (20 mM NaH_2_PO_4_, 0.5 M NaCl, 500 mM Imidazole, pH 7.4). The cartridge was then washed with 20 mM imidazole (98% A, 2% B) and the protein eluted by linear gradient from 2 to 100% buffer B in 10 column volumes. Protein quality was checked by SDS-PAGE and, after dialysis in PBS, the concentration estimated using the Coomassie protein assay and bovine gamma globulin as standard.

### SARS-CoV-2 ELISAs

Upon receipt of patient samples, 100μL aliquots were generated and heat-inactivated at 56°C for 1 hour while shaking at 500rpm. Remaining samples were labeled and stored at −20°C or −80°C. When ready to assess antibody concentration, 20μL of each sample was added to 100μL of 1% non-fat dry milk diluted in PBS + 0.1% Tween 20 (PBS-T) in the first row of 3 pre-blocked and coated Enzyme-linked Immunosorbent Assay (ELISA) plates (Pierce Part #: 15041): one coated with SARS-CoV-2 receptor RBD (2μg/mL), one with N (Sino Biological Part #:40588-V08B) (1μg/mL), and one coated with S1 (Sino Biological Part #: 40591-V08H) (2μg/mL). RBD used to validate the rapid-ELISA prior to serological analysis of patient samples was contributed by David Veesler for distribution through BEI Resources, NIAID, NIH: Vector pcDNA3.1(-) containing the SARS-Related Coronavirus 2, Wuhan-Hu-1 Spike Glycoprotein Receptor Binding Domain (RBD), NR-52422^16^. Samples were diluted five-fold down the plate excluding the final row which served as a negative control for each patient sample. A positive control human monoclonal antibody against an individual antigen was run on each plate to ensure lot-to-lot consistency (human-anti-S1/RBD Sino Biological Part #: A02038 (HC2001), rabbit-anti-N Sino Biological Part #:40143-R001). After sample loading, plates were incubated for 10 minutes at room temperature shaking at 60rpm. Plates were then washed four times with PBS-T. Secondary antibody buffer (100μL of 1 % milk diluted in PBS-T containing 1:500 goat anti-human-IgG-HRP; Invitrogen Part #: 31410) was added immediately following the washing procedure. The plates were incubated for 10 minutes at room temperature shaking at 60rpm. Plates were washed five times with PBS-T. SigmaFast OPD substrate (Sigma Part#: P9187) was prepared in milliQ (18.2MQxcm) water and 100μL was aliquoted into each well. Ten minutes after loading the substrate, 25μL of stop solution (3N HCl) was added to end colorimetric development. The absorbance of the substrate in each well was measured on a Synergy H1 (Biotek) spectrophotomer at 492nm. Antibody concentration was calculated based on area under the curve analyses of A_492_ vs. dilution factor plots for each sample.

### Cytokine quantification

Serum cytokine concentrations of IL-1β, IL-2, IL-4, IL-5, IL-6, IL-8, IL-12, IL-13, IL-18, eotaxin, GM-CSF, GRO-α, IFN-γ, IP-10, MCP-1, MIP-1α, MIP-1β, RANTES, SDF-1α, and TNF-α were assessed using a Human Th1/Th2 Cytokine & Chemokine 20-Plex ProcartaPlex Panel 1 (ThermoFisher Part #: EPX200-12173-901) according to the manufacturer’s instructions. Serum samples (217 samples) were prepared for analysis by heating at 56°C for 1 hour. Samples were then centrifuged at 13,000 x *g* for 2 mins to pellet aggregates. Samples (25μL) were diluted 1:2 with universal assay buffer and incubated at room temperature on an orbital shaker at 500 rpm for 1 hour. Select samples (based on sample quantity) were diluted 1:4 or 1:5 with the universal assay buffer, which was taken into account during analysis. A standard curve was generated using antigen standards provided by the manufacturer. Samples were resuspended in 120 μL wash buffer prior to running on a MAGPIX (Luminex) instrument, and 35 μL was analyzed per samples. Bead counts below 35 were insufficient for analysis and excluded from the analysis.

### CXCL13 quantification

CXCL13 concentration was determined using a Human BLC (CXCL13) ProcartaPlex Simplex Kit (ThermoFisher Part #: EPX01A-12147-901). Plates were coated with magnetic beads according to the manufacturer’s protocol. Plasma samples (25μL, 217 samples) from patients were loaded onto coated plates and shaken for 1 hour at 500rpm at room temperature. Plates were washed 2 times with wash buffer while attached to the magnet before the addition of detection antibody. Samples were shaken (500rpm) for 30 minutes at room temperature to allow for detection antibody binding. Plates were then washed 2 times with wash buffer while attached to the magnet. After washing, 50μL of Streptavidin-PE (SAPE) was added to each well and the plates shaken (500rpm) for 30 minutes at room temperature. Finally, plates were washed 2 times with wash buffer attached to the magnet before the addition of 120μL of reading buffer. Sample aliquots (35μL) were read by the Luminex MagPix instrument with a 35-bead detection limit.

### Principal component and heatmap analysis

Serological data from patients tested for cytokine production and antibody production were pooled into Microsoft Excel and imported to ClustVis^17^. Data were transformed by the ln(x) transformation provided in the webtool and grouped with a 95% confidence interval. Groups were based on patient SARS-CoV-2 status and outcome (survived vs. deceased). Heatmap clustering was based on complete cytokine profile.

### Statistical analyses

Statistical analyses were calculated in GraphPad Prism (version 8.3.0). Comparisons of two conditions were completed using two-tailed Student’s t-tests or Welch’s *t*-tests in cases where standard deviations were different between groups. Statistical significance of multiple variables was assessed using Brown Forsyth and Welch’s one-way ANOVA followed by Tukey’s multiple comparison test. Pearson correlation coefficients and p-values were calculated in GraphPad Prism using the “Correlation” analysis. In all analyses statistical significance was determined to be p<0.05.

## Results

### In-patient anti-SARS-CoV-2 antibody production

Antibody binding target and the timing of the antibody response are critical factors in mediating immunity. We evaluated anti-SARS-CoV-2 antibody production to 3 antigens (RBD, N, and S1) in 82 in-patients (Supplementary Table 1) by developing a novel rapid-ELISA technique. Our rapid-ELISA technology evaluates IgG antibody production to the SARS-CoV-2 RBD, N, and S1 proteins in approximately 1 hour with greater than 99% accuracy (Supplementary Table 2). Our survey of SARS-CoV-2 positive patients demonstrated that antibody (IgG) production to RBD, N, and S1 proteins developed over the first 10 to 20 days post-symptom onset (Figure 1a-c). When comparing antibody production to each antigen, we observed significant IgG production to multiple antigens in the majority of patients tested (219/255 SARS-CoV-2 positive in-patients, 86%), (Figure 1 d-f, Supplementary Figure 1, Supplementary Data File). To better understand the kinetics of the antibody response, we plotted IgG production of every patient over time to RBD, N, or S1. Patients produced IgG against RBD rapidly after symptom onset with the peak IgG response occurring 10 days after symptom onset (Figure 1g). Anti-S1 IgG production escalated over a slightly larger period (13 days, Figure 1 i) and anti-N IgG production was slower than either anti-RBD or anti-S1 antibody production (22 days, Figure 1h). Taken together, these data describe the breadth and timing of the IgG response to SARS-CoV-2 antigens.

**Figure 1.**
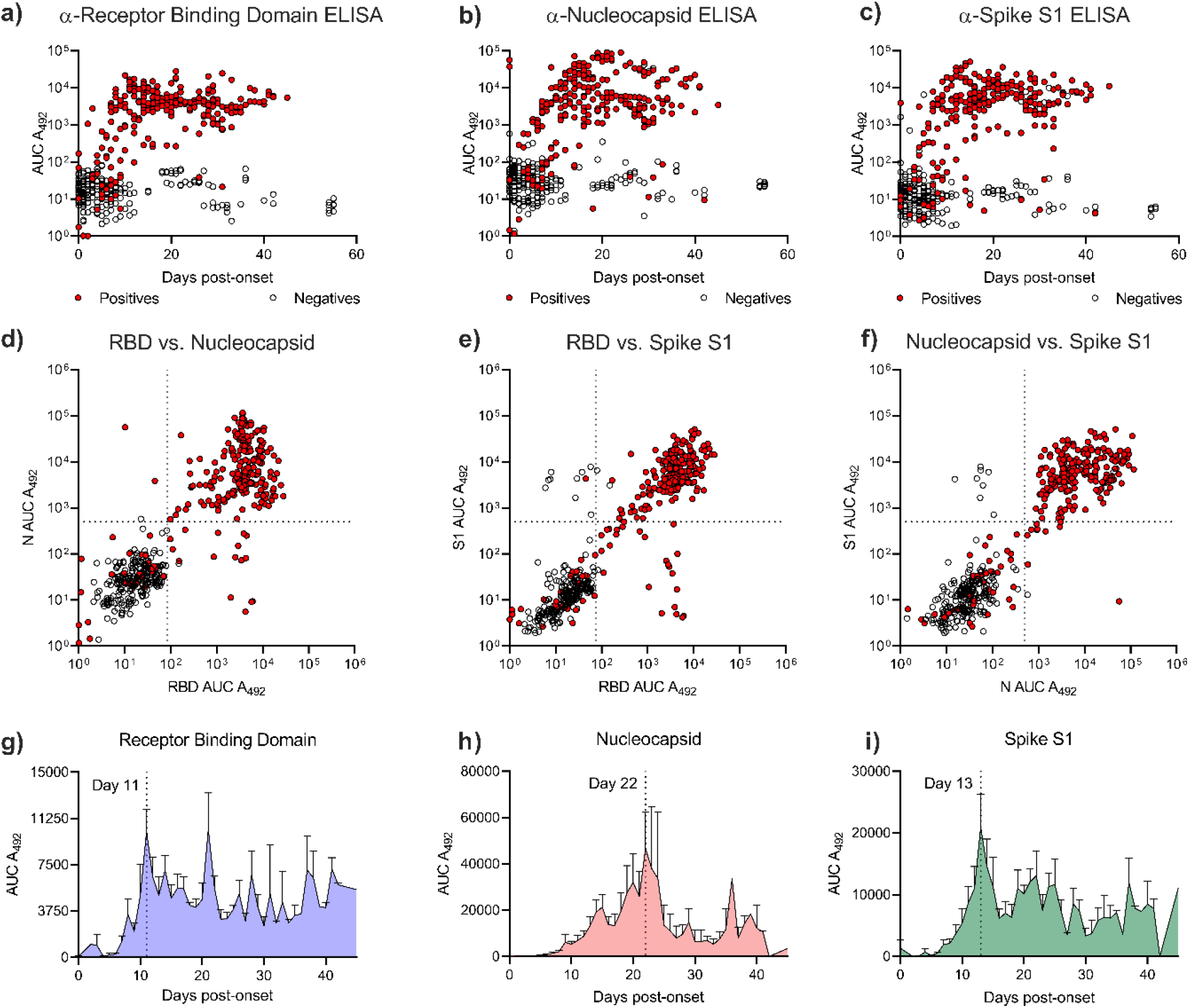
Anti-SARS-CoV-2 IgG response of SARS-CoV-2 in-patients. Antibody (IgG) production to patients that tested PCR positive (red) or negative (clear) for SARS-CoV-2 to RBD (a), N (b), or S1 (c). Correlation of antibody production to RBD vs. N (d) or S1 (e). Correlation of antibody production to N vs. S1 (f). Antibody production of anti-RBD (g), anti-N (h), or anti-S1 (i) antibodies by SARS-CoV-2 positive patients vs. days post SARS-CoV-2 disease onset.

### Antibody production varies depending on patient population

Antibody responses are typically different depending on patient demographics and have implications for population-wide immunity. To understand the anti-SARS-CoV-2 IgG response in different populations, we analyzed patient groups based on sex, patient mortality, blood type, and age against anti-RBD, anti-N, or anti-S1 antibody production. As IgG production is more consistently detectable after ten days post-symptom onset^18,19^, we assessed differences in IgG production beyond ten days post symptom onset. Limiting sample analysis to those greater than ten days post symptom onset did not significantly impact the mean antibody production of the patients (Supplementary Figure 1). Patients who did not survive SARS-CoV-2 hospitalization produced significantly more antibodies to SARS-CoV-2 N than patients that survived infection (Figure 2a). Furthermore, patients that did not survive SARS-CoV-2 infection did not produce different quantities of anti-N antibodies than surviving patients during early infection (Supplementary Figure 2). To accurately assess differences in antibody production independently of disease outcome, we quantified anti-SARS-CoV-2 IgG production in patients who survived infection grouped by biological sex, blood type, and age. We determined that, in our cohort, females significantly produced more anti-S1 IgG than males (Figure 2b). We also observed that blood type was significantly associated with anti-SARS-CoV-2 IgG production (Figure 2c). Blood type B+ patients produced significantly more IgG to RBD and S1 than A+ or O+ patients (Figure 2c) and A+ patients produced the lowest quantities of anti-RBD and anti-S1 IgG. O+ patients produced reduced anti-N IgG relative to A+ or B+ patients. Previous studies have identified that age impacts antibody production to SARS-CoV-2^20,21^. Our study demonstrates that antibody production against RBD or S1 antigens increased with age (Figure 2d). In contrast, antibody production to N increased in patients over 50 years old but did not continue to increase with age after 80 years of age. This is particularly evident when examining Pearson correlations between age and anti-SARS-CoV-2 IgG production for each antigen (Supplementary Figure 3). Overall, these data document a significant impact of patient demographics on anti-SARS-CoV-2 antibody production.

**Figure 2.**
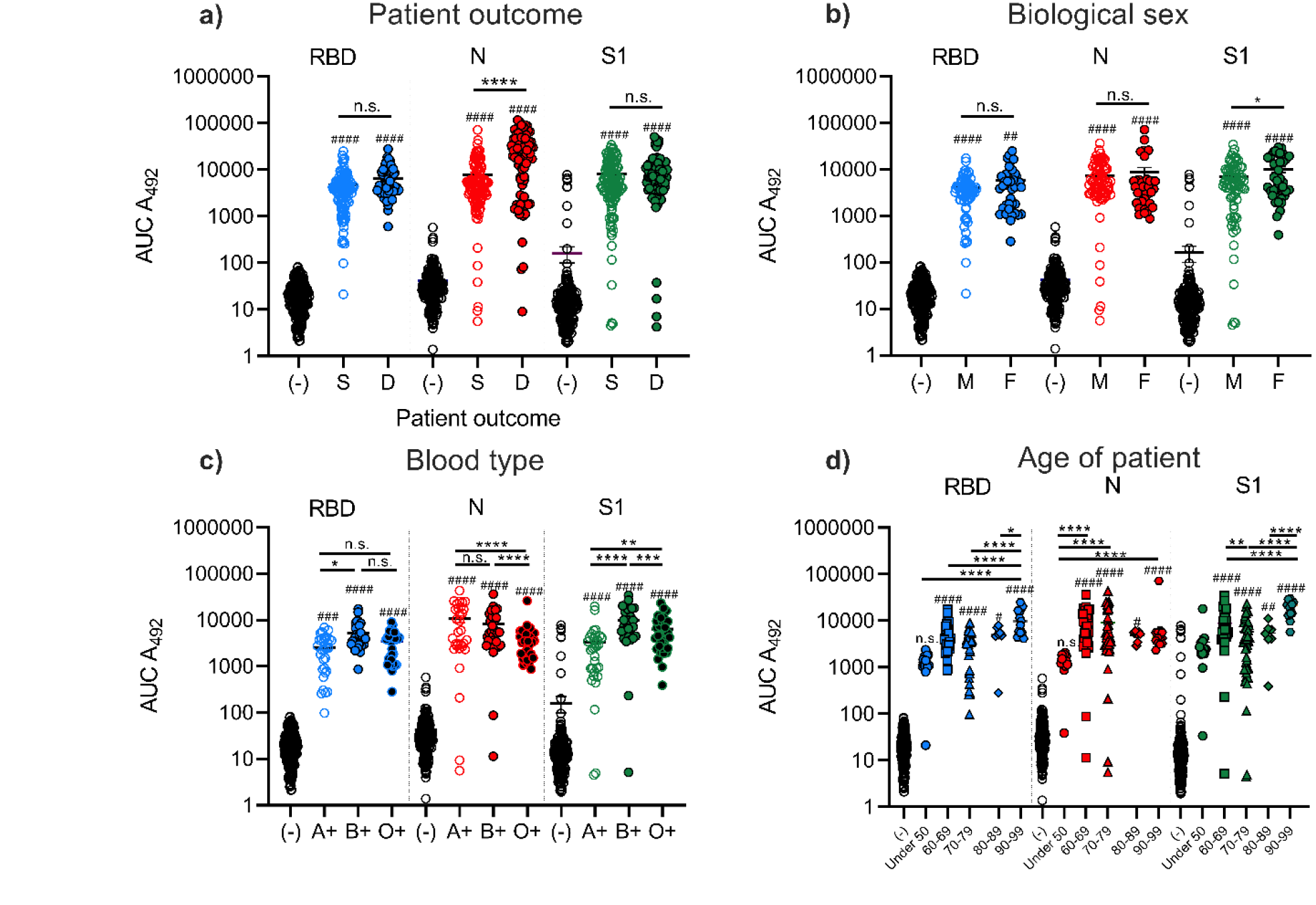
Patient outcome, sex, blood type, and age impact anti-SARS-CoV-2 antibody production. IgG production of patients to RBD, N, and S1 separated based on patients outcome to SARS-CoV-2 infection (a). (S) patients that that survived SARS-CoV-2 infection, (D) did not survive infection, (-) = SARS-CoV_2 negative patients. Anti-SARS-CoV-2 antibody production of surviving patients separated based on sex (b), blood type (c), and age (d). Statistical analysis was completed by one-way ANOVA followed by Sidak’s multiple comparisons test. Significance was assessed between SARS-CoV-2 positive patients and negative patients (#’s) and in between positive patient groups (*’s). ## = p<0.01, ### = p<0.001, #### = p<0.0001, * = p<0.05, ** = p<0.01, *** = p<0.001, **** = p<0.0001, n.s. = not significant.

### Changes in SARS-CoV-2 patient cytokine responses correlate with disease severity

Antibody production represents the antigen-specific response to pathogens but is only one facet of immunity. We examined the broader immunological response to SARS-CoV-2 infection by quantifying the production of cytokines involved in a representative subset of SARS-CoV-2 or healthy patients. SARS-CoV-2 patients exhibited significant increased pro-inflammatory cytokine production (IL-6, IL-8, IL-18) and increased chemotactic cytokine production (IP-10, SDF-1, MIP-1*β*, MCP-1 and eotaxin) relative to non-infected individuals (Figure 3). Of the SARS-CoV-2-infected patients, mortality was associated with increased IL-6, IL-8, IL-18, IP-10, and MCP-1 production. Patients who succumbed to infection also demonstrated intermediate production of SDF-1, MIP-1β, and eotaxin production relative to surviving SARS-CoV-2 patients and healthy individuals. We observed no statistically significant differences in several other measured cytokines between healthy and SARS-CoV-2 positive patients. However we observed that lethal SARS-CoV-2 infection was associated with significantly decreased IL-1β, IL-2, IL-4, IL-12, IL-13, RANTES, TNFα, GROα, and MIP-1α, or increased IFN-γ (Supplementary Figure 4). A representation of a surviving patient’s cytokine profile (Figure 3i) and deceased patient’s profile (Figure 3j) over time are provided. All patient cytokine profiles studied are documented in Supplementary Figure 6. Together, these data demonstrate that SARS-CoV-2 patients exhibit an increased pro-inflammatory and chemotactic response with distinct profiles associated with patient mortality.

**Figure 3.**
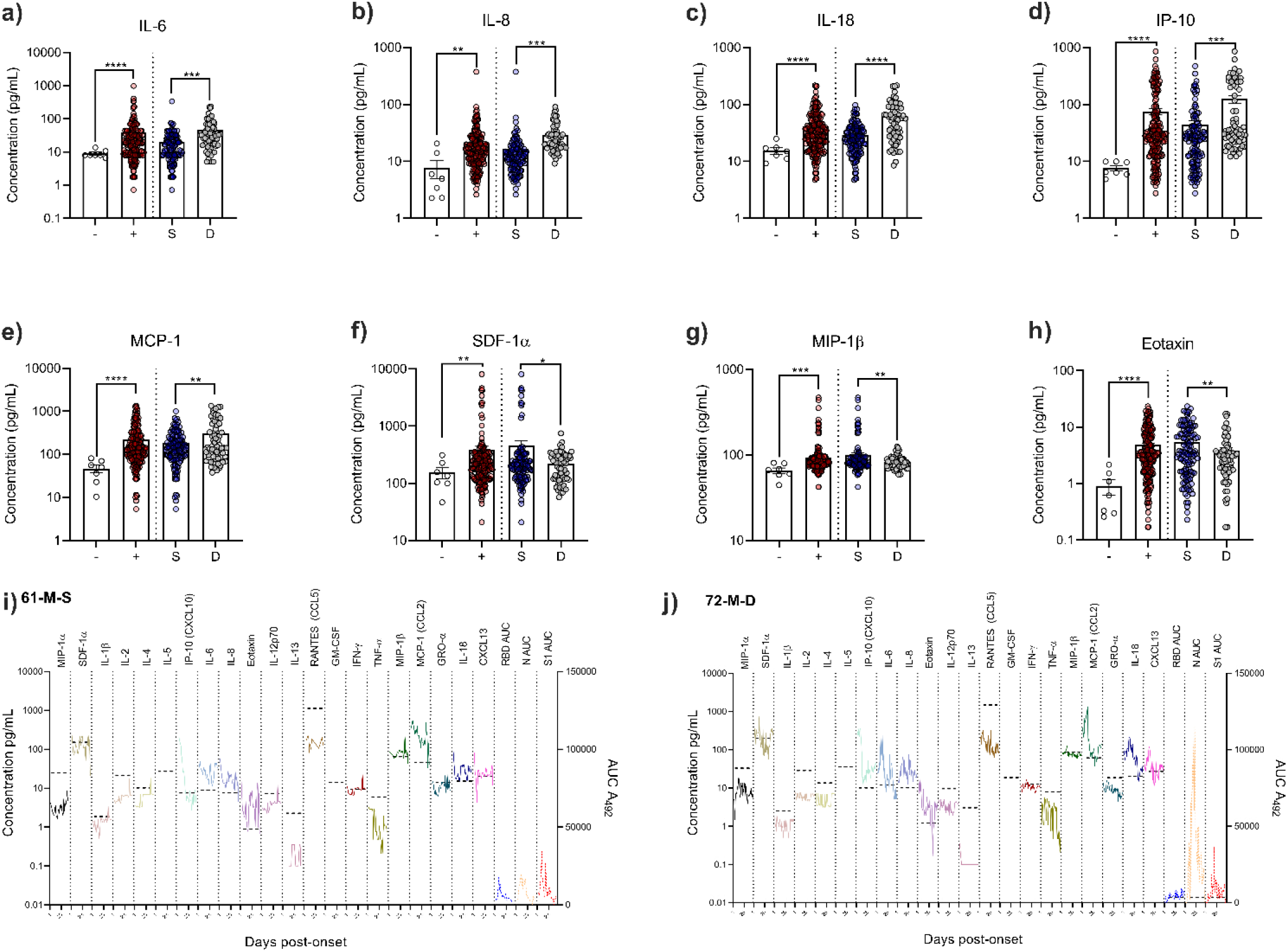
SARS-CoV-2 patient cytokine profile is impacted by disease severity. Concentrations of (a) IL-6, (b) IL-8, (c) IL-18, (d) IP-10, (e) SDF-1α, (f) MIP-1β, (g) MCP-1, or (h) eotaxin were determined by Luminex technology. Full cytokine profiles for a surviving patient (i) or deceased patient (j). – = SARS-CoV_2 negative patients, + = SARS-CoV-2 positive patients, S = SARS-CoV-2+ patients that survived infection, D = SARS-CoV-2+ patients that did not survive infection. Statistical significance was assessed with a two-tailed Welch’s t-test. ** = p<0.01 *** = p<0.001, **** = p<0.0001.

### CXCL13 as a novel predictive tool of lethal SARS-CoV-2 infection

Infectious disease stimulates germinal center formation promoting high-affinity antibody production^8,10,22-24^. This response is critical for eradicating many pathogens. As many SARS-CoV-2 patients produced robust antibody responses to multiple antigens, we hypothesized that germinal center formation would be increased in these patients. To quantify this, we measured the serum concentration of CXCL13, a critical mediator of germinal center formation and a biomarker of this immunological response^8,10,22,23^. We observed that CXCL13 production primarily correlated with peak antibody production to RBD and S1 antigens across SARS-CoV-2 infected patients (Figure 4a-c). Additionally, we observed that there was a significant increase in average production of CXCL13 in positive patients relative to negative SARS-CoV-2 patients. In addition, we discovered that CXCL13 production was significantly increased in patients that did not survive SARS-CoV-2 infection compared to those that did (Figure 4d). When we compared antibody and CXCL13 production based on patient survival over time, we observed that patients who did not survive SARS-CoV-2 infection exhibited a sustained increase in antibody and CXCL13 production relative to surviving patients (Figure 4ef). A full comparison of CXCL13 to anti-SARS-CoV-2 IgGs is provided in Supplementary Figure 5. These results suggest that CXCL13 and intense germinal-center-driven antibody responses are likely associated with lethal SARS-CoV-2 infection.

**Figure 4.**
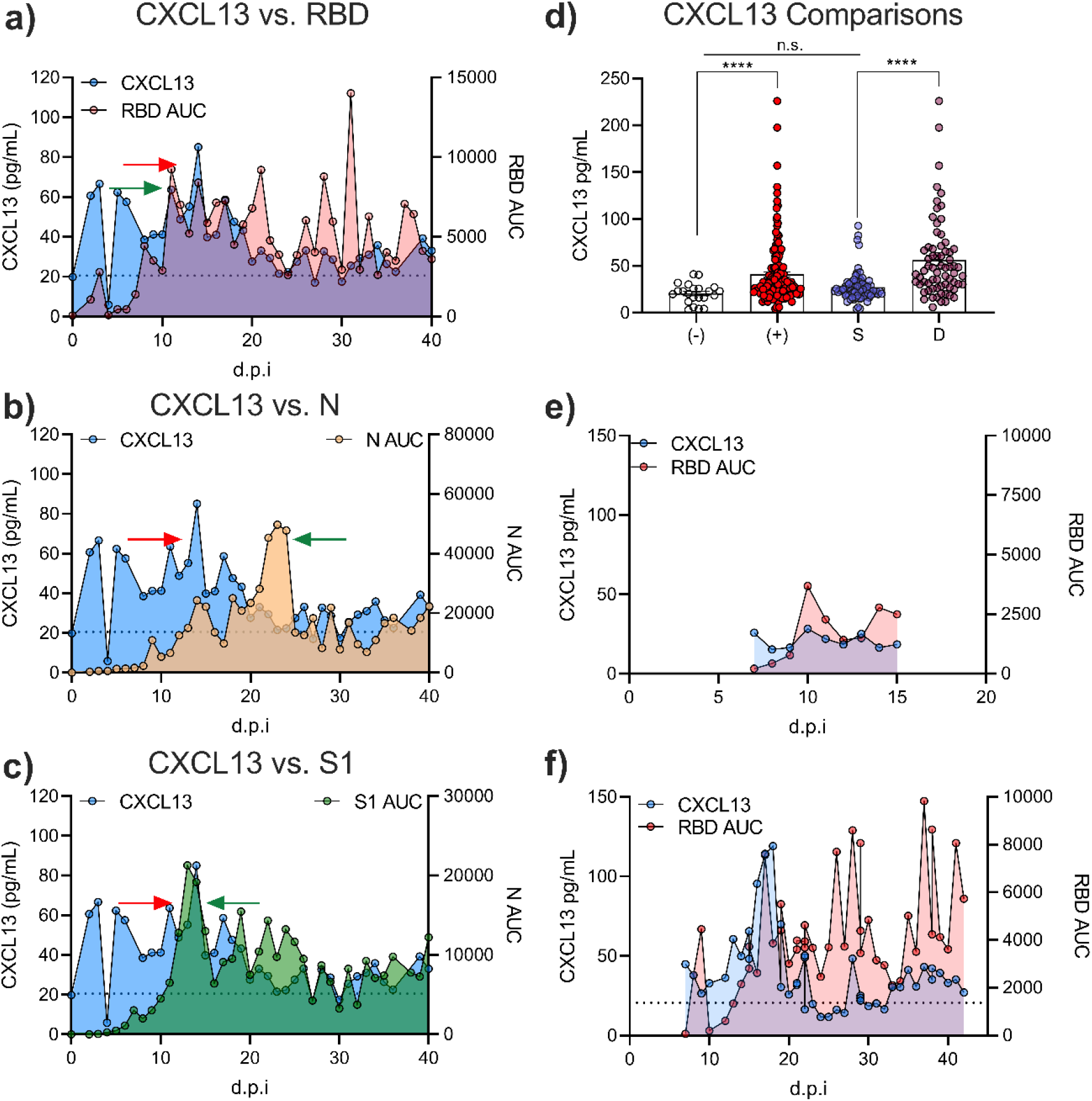
CXCL13 as novel a biomarker for lethal SARS-CoV-2 infection. CXCL13 concentration was measured in SARS-CoV-2 positive and negative patients. CXCL13 production by SARS-CoV-2 production is compared to anti-RBD (a), anti-N (b), or anti-S1 (c) IgG quantity over the course of patient disease. Red arrows represent CXCL13 maxima, and green arrows represent local IgG maxima. CXCL13 production was compared between SARS-CoV-2 negative (-) and positive (+) patients, and SARS-CoV-2 positive survivors (S), or non-survivors (D) (d). Examples of a surviving patient producing low CXCL13 and low anti-RBD IgG response (e) or deceased patient producing high CXCL13 and high anti-RBD IgG response (f). Statistical significance was assessed with a Brown Forsyth and Welch’s one-way ANOVA followed by Tukey’s multiple comparison test. **** = p<0.0001, n.s. = not significant.

## Discussion

Understanding the breadth of the immune response to SARS-CoV-2 infection may be critical to better manage SARS-CoV-2 and prevent it from permeating vulnerable communities on a local and global scale. Initially, we used our rapid-ELISA assay to rapidly assess anti-RBD, anti-N, and anti-S1 antibody production in PCR-positive or PCR-negative SARS-CoV-2 in-patients admitted to a WV hospital. Antibody production against multiple SARS-CoV-2 antigens developed over the course of 20 days post-infection in a manner similar to other studies^19,25-28^. Interestingly, IgG antibody production to N increased over a longer period than antibodies against RBD, or the S1 domain. This could be due to a variety of factors including antigen immunodominance^29,30^, incongruent antigen processing and availability^31,32^, differences in antibody utility and turnover, or prior exposure to similar RBD/S1 antigens of other coronaviruses. Theoretically, as N is not expressed on the viral surface, B cells producing antibodies against this antigen may not be selected for as rapidly as those that are specific to the RBD or S1 antigens and may not possess neutralizing function. As infection worsens, more cells lyse. This may increase the local concentration of free nucleocapsid available for antigen processing and presentation, particularly in lymphoid tissue^33^. In this respect, a more robust antibody response to nucleocapsid later in infection may be due to increased cellular damage. This may initiate a positive feedback loop where infected cells lyse and release nucleocapsid, which induces a less functional anti-nucleocapsid antibody response that fails to alleviate the cell lysis. More evidence is required to support these hypotheses, but these are interesting paradigms to consider in the context of anti-SARS-CoV-2 immunity.

Lethal SARS-CoV-2 infection is significantly correlated with higher antibody production^19,20,26^ and is described further in this study. In analyzing antibody production between patient demographics, it was important to eliminate increased antibody production due to lethal infection as a source of bias. As such, our analyses presented here describe IgG production of SARS-CoV-2 survivors grouped by demographic. There are a multitude of studies reporting differences in IgG production between demographics including: trends in anti-SARS-CoV-2 antibody production between sexes^20,21,34-36^, a correlation of genetically encoded blood type with SARS-CoV-2 immunity^37^, and variability in antibody production in the aging population^20,21^. From these prior studies and others^38,39^ it is known that biological sex can impact antibody production during infection. We observed this phenomenon when quantifying sex specific anti-S1 IgG production. The anti-viral response is mediated in part by Toll-like receptors which are differentially regulated between the sexes^40,41^. A higher frequency of anti-S1 IgG in females would suggest an increased neutralizing response to the virus which has not been thoroughly evaluated to-date. Our data exhibited a modest difference in antibody production between sexes. As a result, we do not consider biological sex to be a major contributor to anti-SARS-CoV-2 antibody production.

It is documented that red blood cell phenotypes can influence microbial pathogenesis as antigens can function as receptors and/or co-receptors for pathogenic organisms^42^. Historically, an association was identified between ABO type and pathogen infectivity during the SARS-CoV Hong Kong hospital outbreak in 2003; during that outbreak a small cohort of type-O healthcare workers showed significantly decreased odds of infection relative to health care workers with other blood types^42,43^. An additional study demonstrated that antibodies against the A blood type antigen can inhibit SARS-CoV spike protein binding to ACE2^44^. Although the underlying mechanism relating blood type to SARS-CoV-2 pathogenesis remains unclear, it appears there may be a relationship between ABO blood type and coronavirus infection. Recent data identified the 9q34.2 locus (ABO blood group locus) as potentially involved in susceptibility to COVID-19 respiratory failure with evidence that type A phenotypes are at higher risk while type O phenotypes are partially protected^45^. The data generated in these studies show an interesting pattern that may reinforce blood type related outcomes in severe disease due to a previously unreported association to the level and type of antibody response. As seen in Figure 2c, the relative quantity of anti-RBD and anti-S1 antibodies was highest among type-O and -B individuals and lowest in type-A individuals while the opposite is true of anti-N antibodies. This is further accentuated by evaluating the ratio of anti-RBD or anti-S1 versus anti-N in our patient cohort which shows that higher N:RBD or N:S1 ratios are associated with poor prognosis (Supplementary Figure 5). It is plausible that type-A individuals may have a misdirected humoral response due to antigenic homology between N-acetyl-galactosamine sugar moieties on the A antigen and Spike protein resulting in molecular mimicry. This would result in type-O and -B individuals registering more Spike protein epitopes as foreign and eliciting a more robust humoral response; in turn, this putative mechanism could reduce infectious dose and decrease the risk of mortality. Further studies evaluating physiologic modifications of Spike protein and its antigenic moieties would help support or disprove this theory. As the conclusions from these observations are currently theoretical, a more extensive review of comorbid conditions – with a multivariate analysis and estimations of associated odds ratios – may reveal other associations outside of blood type.

The aging process is associated with decreased T-cell functionality^46^, resulting in hyperactive B-cell proliferation that does not confer immunity^47^. We discovered that older patients typically produced more antibodies to RBD and S1 than younger patients. The lack of increase in antibody production to nucleocapsid in the elderly may be a function of antigen availability. To speculate, if elderly patients have higher viral loads due to decreased remediation of virus this would increase the relative abundance of surface exposed antigens (RBD and S1), but not necessarily hidden antigens (N). Increased antibody production would therefore predominantly occur to RBD and S1, and not N. Other challenges are associated with studying this population including co-presentations of multiple diseases which complicates this analysis. Regardless, our study has identified several patient demographics associated with differences in the anti-SARS-CoV-2 antibody response.

The anti-viral immune response depends on a variety of signaling pathways mediated by cytokines and chemokines. Many of the pro-inflammatory cytokines associated with the anti-viral response are upregulated in patients with lethal SARS-CoV-2 infection in our study. IL-6, IL-8, and IL-18 are known pro-inflammatory cytokines that aid in the antiviral response and have been identified in other studies of SARS-CoV-2 patients^2,48-52^. These cytokines are considered part of the “cytokine storm” notorious for inducing localized tissue damage, which may explain the relative increase of these cytokines in deceased patients. In general, the production of these cytokines was similar in SARS-CoV-2 patients to individuals with other lethal viral infections^53-55^.

Chemotaxis is another critical component of antiviral immunity and several chemotactic mediators were increased in patients from our study. IP-10 is a chemotactic agent that was increased ten-fold in SARS-CoV-2 patients and even more so in deceased SARS-CoV-2 patients. IP-10 is protective in SARS infection^56,57^ suggesting that this may be a critical component of anti-SARS-CoV-2 immunity. In a broader sense, this chemotactic response likely functions by inducing chemotaxis of phagocytic immune cells and activated T cells similar to infection with other viral infections^58^. Several other chemoattractive mediators with similar function were upregulated in SARS-CoV-2 patients revealing a systemic increase in leukocyte recruitment (Figure 3, Supplementary Figure 4). Two functional outliers in this analysis were SDF-1 and eotaxin, which are particularly interesting chemokines with broader functional capabilities. SDF-1 is a potent chemoattractant for leukocytes^59^, but has also been implicated in cardiac stress signaling and repair mechanisms^60-62^. Given increasing reports of cardiac disease concurrent with, or following SARS-CoV-2 infection^63,64^, the increase in SDF-1 may be an indirect indicator of cardiac distress. Separately, we discovered increased eotaxin production in SARS-CoV-2 patients. Eotaxin was increased or similar to healthy patients during SARS-CoV-2 infection in other studies^49,65^. Eotaxin is typically involved in eosinophil recruitment, which can result in pulmonary damage^66^. This chemokine is upregulated during viral infection^67^, and can inhibit certain viral infections, such as HIV^68^. As patients who survived infection produced significantly more eotaxin than patients with lethal infection, it is possible that eotaxin provides a double-edged function in SARS-CoV-2 immunity.

Surprisingly, we did not observe changes in production of a number of other cytokines that are involved in the general anti-viral response (i.e. TNF-α; Supplementary Figures 4 and 6). Although this was the case, the noticeable decreases in IL-1β, IL-2, IL-4, IL-12, IL-13, RANTES, TNFα, GROα, and MIP-1α observed in patients with lethal SARS-CoV-2 infection, suggest that lethal infection results in an exhausted immune response^69,70^. When considering the overall cytokine response to SARS-CoV-2 in conjunction with the anti-SARS-CoV-2 antibody response, it is clear that distinct phenotypic clusters of healthy patients, surviving patients, and patients with lethally-infected (Figure 5). In this respect, these analyses paint a more definitive picture of the anti-SARS-CoV-2 cytokine response. Despite the significant results of this study, these data should be evaluated in broader context as patient demographics, treatment plan, and course of infection likely play a role in differences in cytokine production between patients and studies. Large-scale analyses of cytokine production on a population-wide scale would likely be necessary to fully understand the cytokine profile of anti-SARS-CoV-2 immunity.

**Figure 5.**
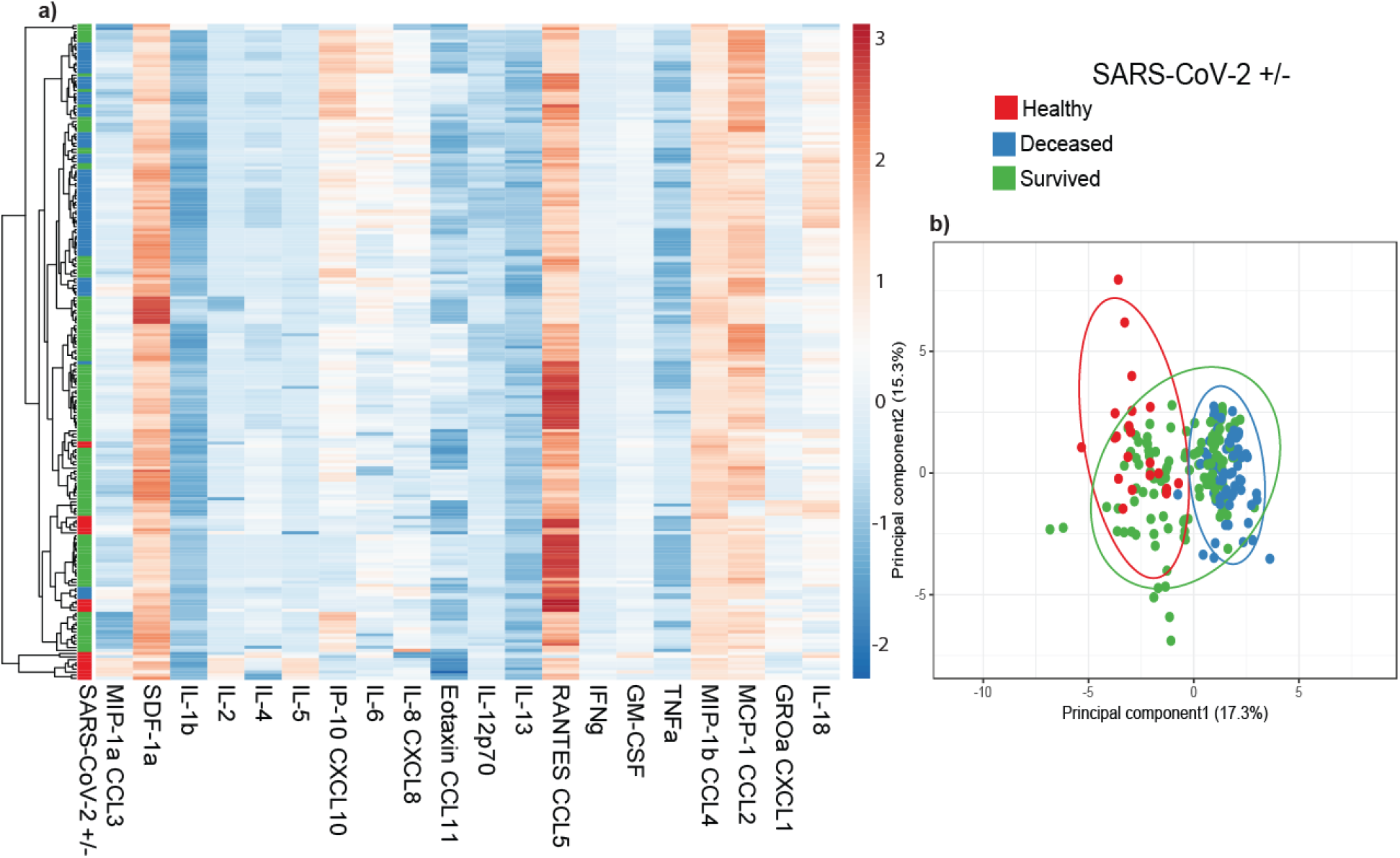
Principal component analysis of anti-SARS-CoV-2 immunological responses. Heatmap (a) and principal (b) component analysis of all patient samples including 20 cytokine concentrations clustered using ClustVis^17^.

Antibody maturation signaling has not been investigated in the context of SARS-CoV-2. We assessed the activity of the antibody maturation pathway by measuring CXCL13 concentrations in the serum of SARS-CoV-2 patients. Increased CXCL13 in SARS-CoV-2 patients may indicate heightened germinal center activity^8^ and affinity maturation of anti-SARS-CoV-2 antibodies. The significant increase of CXCL13 in patients with lethal disease suggests this may be an emergency response to uncontrolled infection. It is possible that sustained infection stimulates increased antibody affinity maturation that is unable to keep pace with viral replication and the cytokine storm. In this sense, CXCL13 could be used as a marker of SARS-CoV-2 disease severity. There is a precedent for the utility of CXCL13 as a biomarker that is predictive of immune activation during HIV exposure^8,9,24^. This adds credibility and feasibility for this utility, but further studies are required to validate this approach. We have provided a schematic of how the CXCL13 response interplays with our other observations of SARS-CoV-2 immunity in Figure 6.

**Figure 6.**
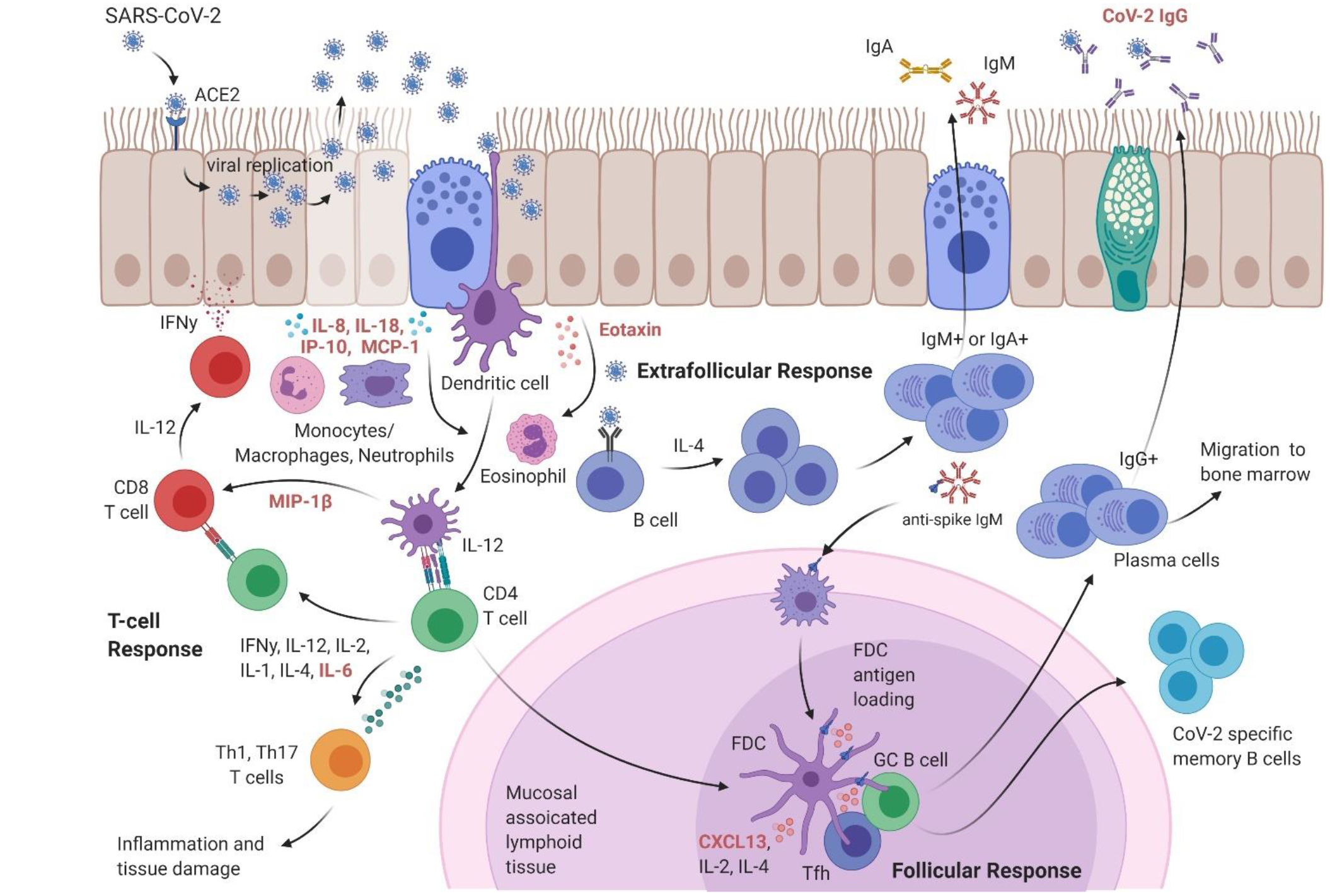
Overview of SARS-CoV-2 immunity. A schematic of the findings provided in this study (increased immunological markers highlighted in red) in the context of anti-SARS-CoV-2 immunity.

To summarize, this study provides insight into the breadth of the immunological response against SARS-CoV-2. We demonstrated increasing antibody production to multiple SARS-CoV-2 antigens over the first ten days of infection using a rapid-ELISA assay. Our results exhibit that patient mortality, sex, blood type, and age impact antibody production to SARS-CoV-2, adding to what is known about SARS-CoV-2 pathogenesis. Furthermore, lethal SARS-CoV-2 infection triggers a pro-inflammatory cytokine response, in combination with the secretion of several chemotactic agents. Interestingly, patients with lethal SARS-CoV-2 disease exhibited divergent cytokine production compared to patients with non-lethal disease. Finally, we discovered that a marker of germinal center activity (CXCL13) is upregulated in SARS-CoV-2 patients, and that this upregulation is amplified in lethal disease. Ultimately, these studies help to elucidate the interplay between immunological responses to SARS-CoV-2 and identify a potential novel biomarker of COVID-19 severity.

## Data Availability

All data in this manuscript is provided in associated supplementary files.

## Acknowledgements

AMH, TK, and FHD designed the experiments. TK, JMK and BJH compiled and transferred patient data. AMH and BRR ran rapid-ELISA assays of patient samples. AMH, MAW, and MAD performed CXCL13 quantification assays. MAW, and MAD performed 20-plex cytokine assays. PF produced RBD used in this study. AMH analyzed and compiled assay data and figures. All authors took part in writing and editing the manuscript. We would like to thank BEI Resources for providing the following reagents (NR-52422). We would finally like to express our gratitude to Drs. Laura Gibson and Clay Marsh for enabling this research during the global pandemic.

